# A Quantitative Framework for Evaluating the Performance of Algorithm-Directed Whole-Population Remote Patient Monitoring for Type 1 Diabetes Care

**DOI:** 10.1101/2025.02.03.25321621

**Authors:** Jamie Kurtzig, Johannes O. Ferstad, Paul Dupenloup, Dessi P. Zaharieva, David M. Maahs, Priya Prahalad, Ramesh Johari, Franziska K. Bishop, Ananta Addala, David Scheinker

## Abstract

**Introduction:** Clinics continue to adopt remote patient monitoring for type 1 diabetes (T1D) and care models shaped by algorithmic CGM data analysis. No clinic-facing quantitative framework currently exists to track the impact of such algorithm-directed care on patient outcomes and clinical workload. The Teamwork, Targets, Technology, and Tight Control (4T) Study provides precision, whole-population care enabled by algorithms that use continuous glucose monitoring (CGM) data to direct clinician attention to patients with deteriorating glucose management.

**Methods:** We used data from the 4T Pilot (n=133) and 4T Study 1 (n=135), in which algorithms use CGM data to identify youth with T1D meeting criteria for clinical review and potential clinician contact. Through iterative data analysis and interviews with diabetes educators and clinicians, we identified metrics for reviewing and revising clinical workloads, glucose management, and timeliness of care. For each metric, we developed an interactive dashboard to provide clinical and administrative leaders with an overview of the program.

**Results:** The metrics to track clinical workload were the total number of youths: (1) in the program, (2) in each study, and (3) cared for by each clinician. The metrics to track glucose management were the number of youths meeting each criterion for review: (4) in total, (5) for each clinician, and (6) for each study. The metric to track timeliness of care was (7) the number of days since meeting criteria for clinical review. When presented at weekly program leadership meetings, the metrics facilitated data-driven decision making about clinical and operational components of the program.

**Conclusion:** We propose a novel quantitative framework for diabetes care teams to supervise and enhance algorithm-directed whole-population T1D care. As the role of algorithms grows in directing clinical effort and prioritizing patients for care, this framework may help clinics track clinical workload, patient outcomes, and the timeliness of care.

## Introduction

Remote patient monitoring (RPM) programs for type 1 diabetes (T1D) care often use data provided by continuous glucose monitoring (CGM) systems.^1–4^ RPM for T1D has been shown to improve HbA1c, quality of life, and glucose metrics in a variety of healthcare settings.^5–7^ Numerous centers report the use of RPM for T1D care, and many include some level of algorithmic support in identifying patients for prioritization and clinical review or contact.^8–12^ In the Teamwork, Targets, Technology, and Tight Control (4T) Study 1 cohort, care directed based on the analysis of CGM data equitably reduced median HbA1c to 6.58% at 12 months after diagnosis.^12^ The growing use of CGM is driving the demand for algorithms to manage data and inform care delivery, increasing the complexity of clinic operations.^13^

To our knowledge, no clinician-facing quantitative framework is available to track how algorithm-directed care impacts clinical workload, patient glucose management, and timeliness of care. Such quantitative frameworks may be helpful for clinics that: remotely access patient data; provide RPM-based care; and employ algorithms to direct care delivery by, for example, identifying or prioritizing patients requiring care. Although our work focuses on RPM in a pediatric T1D population, these concepts may translate to other chronic health conditions in which algorithms support and guide clinical care.^14–16^

The Stanford 4T Study whole-population RPM tool, Timely Interventions for Diabetes Excellence (TIDE), uses CGM data to prioritize patients with T1D for personalized review and contact by the Certified Diabetes Care and Education Specialist (CDCES).^13,17^ The 4T Study and the use of TIDE have been associated with significant, equitable improvements in glycemic outcomes, as well as reduced CDCES workload and burden.^17–22^ The aim of this work is to develop a set of clinical and operational metrics for an organization to monitor and improve population-level, data-driven T1D care.

## Methods

### Setting

The 4T Study aims to improve care for newly diagnosed youth with T1D in Stanford’s Pediatric Endocrinology clinic. The study was approved by Stanford’s Institutional Review Board (clinicaltrials.gov: NCT03968055, NCT04336969). Participants ≥ 18 years of age or legal guardians of minors provided informed consent, while youth ≥ 7 years of age provided assent prior to study initiation. Participants in the 4T Study were between the ages of 6 months and 21 years and started on CGM (Dexcom G6; Dexcom Inc., San Diego, CA) within the first month of T1D diagnosis. A summary of the program is provided below, and details of the program have been published.^12^

The 4T Study team performed asynchronous remote patient monitoring of CGM data from participants using an algorithm-enabled care platform, Timely Interventions for Diabetes Excellence (TIDE). The 4T Study includes the Pilot 4T study and 4T Study 1, as well as long-term follow-ups for each study. The long-term follow-up programs assess the ongoing impact of the Pilot 4T and Study 1 over an extended period. The Pilot 4T Study (n=135) enrolled participants from July 2018 to June 2020.^10,18^ Participants diagnosed between March 2019 and January 2020 were initially offered non-TIDE RPM (e.g. Dexcom Clarity), which provided individual T1D insights rather than population-wide overviews like TIDE. These participants transitioned to TIDE when it was launched in January 2020. Participants diagnosed during and after January 2020 were enrolled into TIDE for RPM. 4T Study 1 (n=133) enrolled participants from 2020 to 2022 and focused on initiating CGM use within the first 30 days after T1D diagnosis.

In both the Pilot 4T and Study 1, participant data were initially reviewed weekly for the first year after T1D diagnosis and then monthly thereafter.^23^ After the conclusion of each study, participants had the option to participate in an ongoing long-term follow-up program with monthly data reviews conducted by a CDCES.

### RPM Platform

Participants uploaded their CGM data via a personal smart device or study-provided iPod Touch (Apple Inc., Cupertino, CA) to Dexcom Clarity. TIDE automatically pulled CGM data from the 4T Study’s Dexcom Clarity clinic account using a Python script, analyzed the data, and displayed results in an interactive data visualization dashboard called Tableau. Guided by CGM clinical consensus metrics, the TIDE algorithm ranked participants based on the urgency of data review and contact by a CDCES.^20,20,23,24^

Each week, the TIDE platform tracked whether participants met the following clinical categories: spending less than 65% of CGM time in range (TIR; 70-180 mg/dL), over 1% time below range (TBR) level 2 (below 55 mg/dL), a drop in TIR over 15% points from the previous week, over 4% TBR level 1 (below 70 mg/dL), over 50% missing CGM data, or meeting clinical targets.^25^ Meeting targets refers to all participants that do not meet any of the other criteria (i.e., they have > 65% TIR, < 1% TBR level 2, < 15% drop in TIR, < 4% TBR level 1, and < 50% missing CGM data). Some metrics were updated between the Pilot Study and Study 1; for example, the TIR target was changed from 65% to 70%, changing the criteria flagged.

The CDCES team members used TIDE to view a list of participants, ranked by their clinical risk likelihood as defined by the CGM metrics listed above. CDCES team members then took appropriate follow-up steps if needed, such as sending secure messages with care recommendations to participants through the Electronic Health Record (EHR).

### Metrics and Key Performance Indicators (KPIs)

A metric is a quantitative measure of an aspect of a care process or the patient population. A key performance indicator (KPI) combines metrics into a high-level measure or visualization that reflects strategic goals and provides insights into the overall success and performance of an organization. The KPIs presented in this paper include clinical workload, glucose management, and timeliness of care.

### Developing a Quantitative Framework for Clinic Management

In 2022, we began developing a standardized approach to monitor the clinical workload, patient glucose management, and timeliness of care associated with the use of TIDE. Numerous ad-hoc metrics were developed and measured based on the experiences of the 4T Study team, CDCES team, and administrative team.^20^ The care team evaluated each proposed metric. We discarded metrics that were not clearly interpretable, did not translate into a useful KPI, or did not inform a clinical or operational decision. The next stage was an analytical approach to refine the metrics based on an analysis of the data generated, structured interviews with the CDCES team, and discussions with the research and clinical staff. In interviews, the stated goals of the process were to establish a more robust and comprehensive set of metrics for monitoring clinical workload, patient glucose management, and timeliness of care in the presence of algorithm-directed care.

The above process of developing, evaluating, and soliciting feedback was used iteratively to develop metrics and KPIs based on these metrics. If a metric was interpretable and had the potential to drive a specific decision, we developed multiple visual representations. We shared these visualizations with the CDCES team and the clinicians in the 4T Study and asked them to identify specific decisions that could be informed by the metric. After receiving feedback, we refined the definition and visual presentation of each metric and eliminated from further consideration metrics that did not inform specific decisions. We asked the care team if each metric was appropriate and how it would be interpreted. Along with the definition of each finalized metric, we provided the KPIs that the metric is used to generate, and illustrative examples of the decisions it supports.

### Visualization of Metrics

We developed an instantiation of this framework with clinic-specific metrics deployed in an interactive dashboard engineered to streamline the monitoring and visualization of the entire program. We note that participants were enrolled in and departed from the 4T Study continuously; thus, the number of participants shown at any time in the visualizations is less than the total study population.

### Practical Use of the Dashboard for the Visualization of Metrics

At the weekly 4T study team meeting with clinical and administrative 4T team members, the dashboard was presented, and the metrics were discussed to facilitate an overview of the program. Feedback from the 4T team on potential improvements to the metrics and associated visualizations were incorporated. All visualizations developed in Tableau were double-checked with manual calculations.

### Obtaining Feedback

We evaluated the perceived utility of this framework through interviews with the care team and discussions at the weekly 4T team meeting. The interviews were designed to collect qualitative insights into the clinical team’s firsthand experiences, perceptions, and suggestions regarding the framework’s implementation and impact. We ensured that a diverse range of perspectives were represented, including CDCES team members, clinical research coordinators, clinicians, engineers, and research staff.

## Results

Of the 268 participants in the 4T Pilot and Study 1, a total of 222 were eligible for RPM and were included in the KPI framework (Table 1). In the Pilot Study, 46 patients were ineligible, as TIDE had not yet been developed.^10^ The metrics were calculated from data continuously pulled from servers on each participant’s data completeness, the study they were enrolled in, the most recent week they were shown in the TIDE dashboard, their CGM data, and the CDCES responsible for reviewing their data.

**Table 1.**
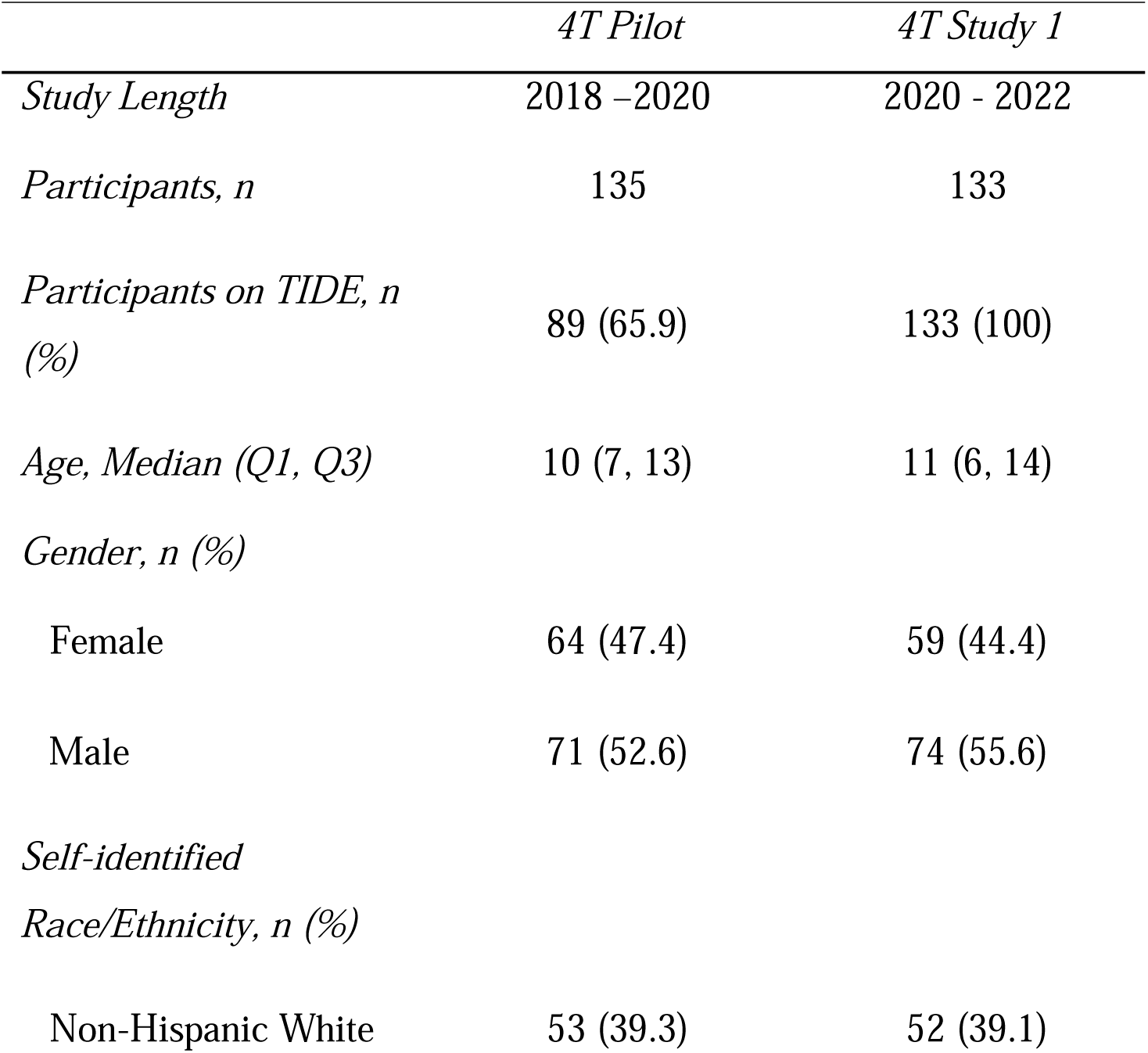
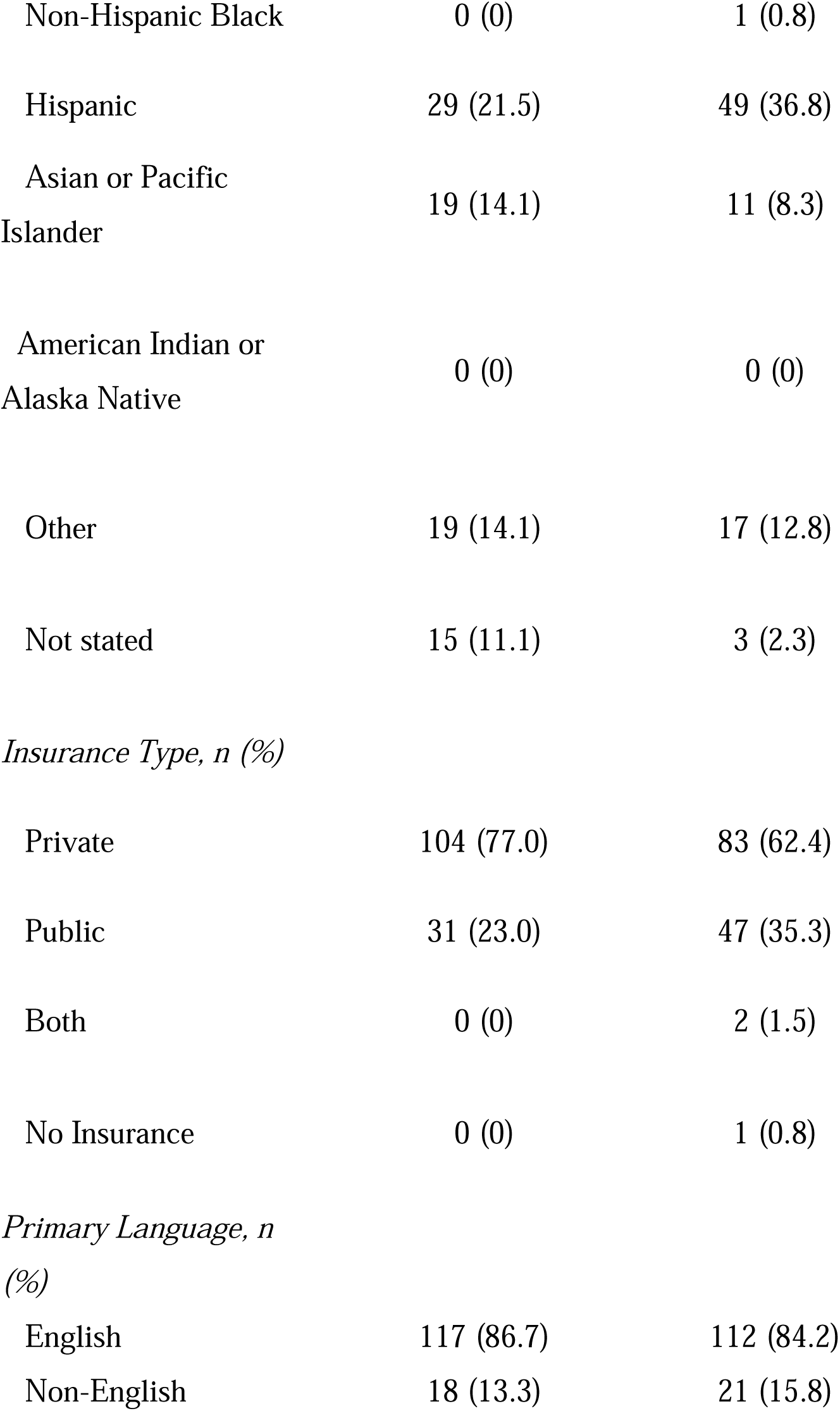
Participant demographics for the 4T Pilot and 4T Study 1.

To monitor clinical workload, the following metrics were identified: (1) the number of youths in the 4T program, (2) in each study, and (3) cared for by each CDCES. For tracking participant glucose management, the metrics include the number of youths meeting each criterion for clinical review, both (4) overall and for each (5) CDCES and (6) study. Initially, the metric for tracking the timeliness of care was the number of days since each participant’s last day meeting criteria for clinical review. This was later revised to (7) include only participants who have not been seen in over 20 days and rank in the top 25% of patients with the longest time since meeting criteria. Feedback from all care team members highlighted these seven metrics as essential for monitoring clinical workload, patient glucose management, and timeliness of care (Table 2). The detailed definitions of each metric are provided in the appendix. Each metric is associated with a KPI displayed in an interactive figure with various filters and categorizations (Table 2).

**Table 2.**
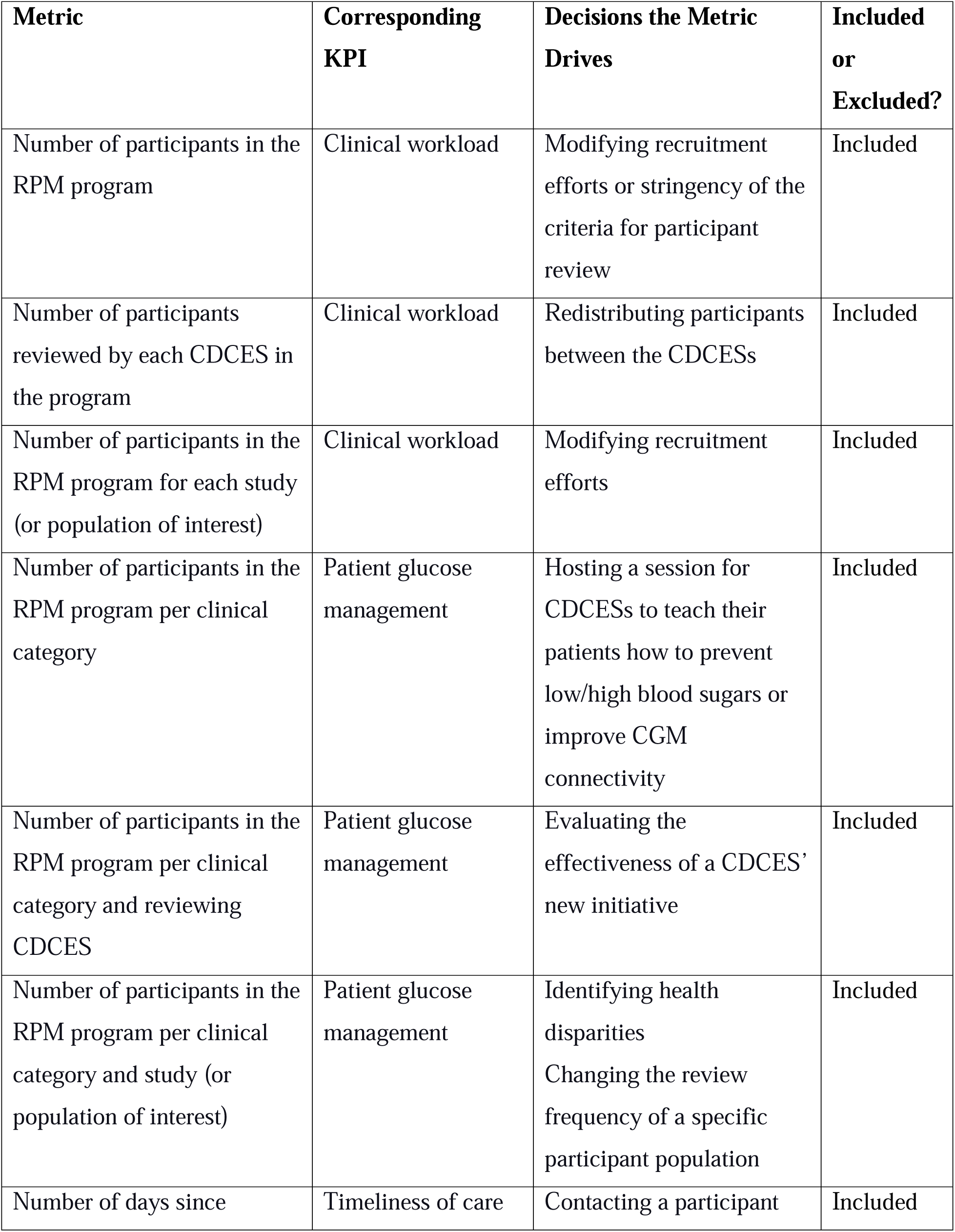
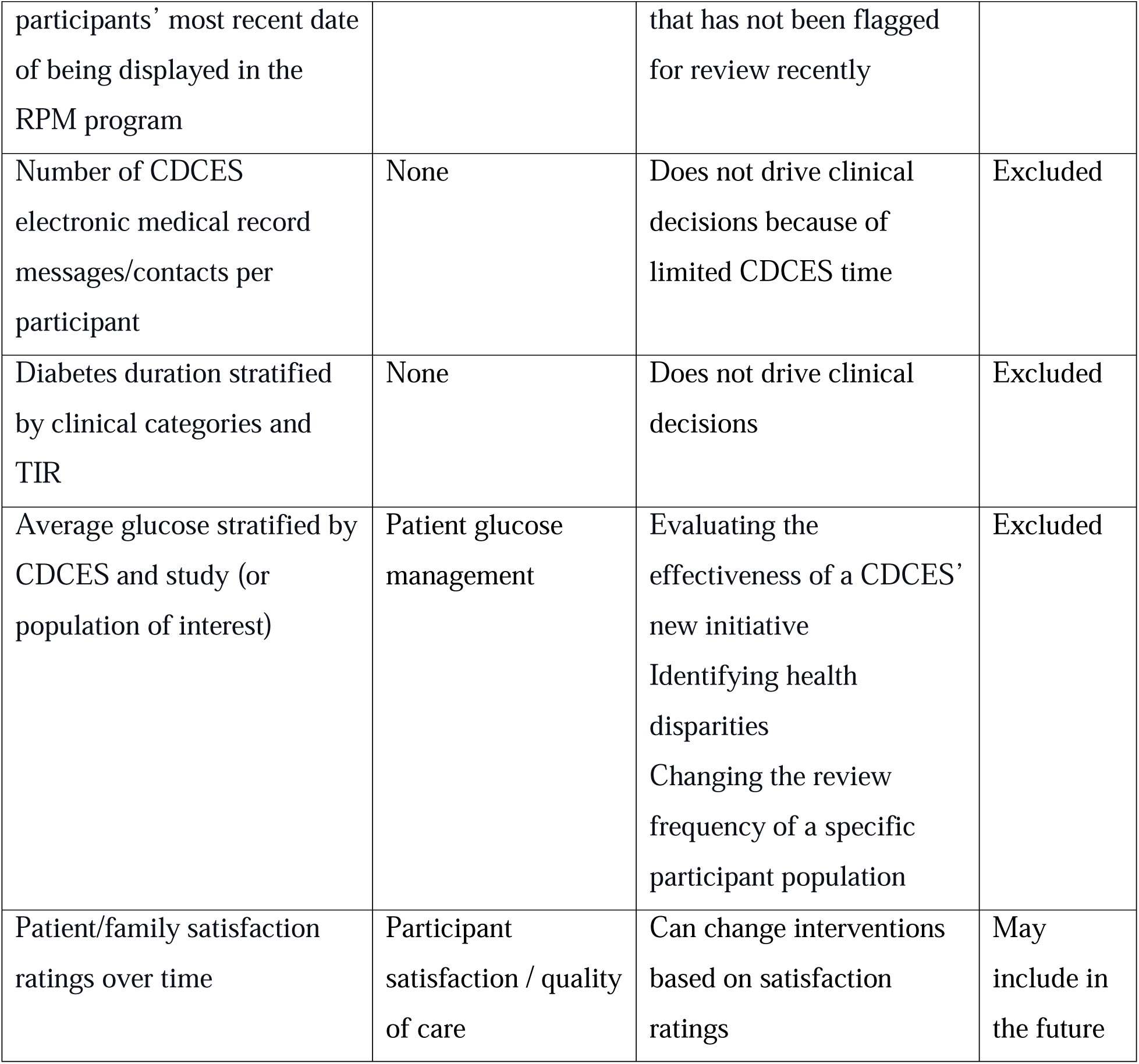
Included and excluded metrics, the corresponding key performance indicators (KPIs), and the decisions the metrics drive.

To visualize each metric related to clinical workload, we used different colors to represent the number of patients: requiring review, meeting targets, and missing >50% CGM data. For each of those three criteria, this was reported as the number of distinct patient IDs.

In the visualization of metrics related to patient outcomes, we used different colors to represent the number of patients in each clinical category. The clinical categories are less than 65% TIR, over 1% TBR level 2, over 4% TBR level 1, over a 15% drop in TIR, and less than 50% CGM wear time. For each clinical category, this is defined as the number of distinct patient IDs of participants flagged.

For the visualization of the metric related to timeliness of care, we displayed participant information only if more than 20 days had passed since the participant was last shown in the TIDE dashboard, and the participant was in the highest quartile for delayed care. The threshold of 20 days was selected based on a retrospective analysis of historical TIDE data, which revealed that the 75th percentile of time between participant dashboard appearances was 20 days.

Throughout this process, three metric categories were discarded because they were difficult to interpret or did not support a clinical or operational decision. The first were measures of the number of CDCES messages/contacts through the EHR, which included metrics such as overall message count, contact count, TIR by message count, CGM time worn by message count, patients contacted versus suggested for contact, and patient response rate to clinician messages (Table 2). Due to limited CDCES time, it was decided that these metrics would not drive clinical decisions. Second were measures of how diabetes duration may impact the clinical categories and CGM metrics, as well as the average diabetes duration for each of the CDCES’ patients (Table 2). These were ultimately excluded since they would not drive day-to-day clinical or operational decisions and would be more appropriate for investigation in retrospective analyses. Third were measures of participants’ average glucose (Table 2). These were excluded since TIR is highly correlated and considered a more comprehensive measure.^25,26^

For all metrics and visualizations presented, gaps or valleys in the data occur when, primarily due to the cadence of reviews, fewer CDCESs used TIDE during that period.

### Metrics and Visualizations for Clinical Workload

The first metric was the total number of participants shown in the TIDE program. By monitoring the number of participants shown in TIDE, this metric allowed for the tracking of population statistics over time. The visualization of the metric selected by the CDCES team included the total number of participants in the program over time, segmented by those that did and did not require review (Figure 1). The periodicity in the figure may be explained by the unequal number of participants assigned to each week (Figure 1). The repetitive patterns in the data are due to the cadence of CDCES reviews. For example, 4T Pilot initially reviewed participants weekly but switched to monthly after 1 year of diagnosis, which contributes to the large decrease in participants seen around week 40 of 2022. One decision potentially driven by this metric was investigating recruitment efforts or participant withdrawal rates if the metric were to be unexpectedly low or high. This metric was also used to investigate the stringency of the criteria for participant review if the numbers of participants being reviewed were too high or too low.

**Figure 1.**
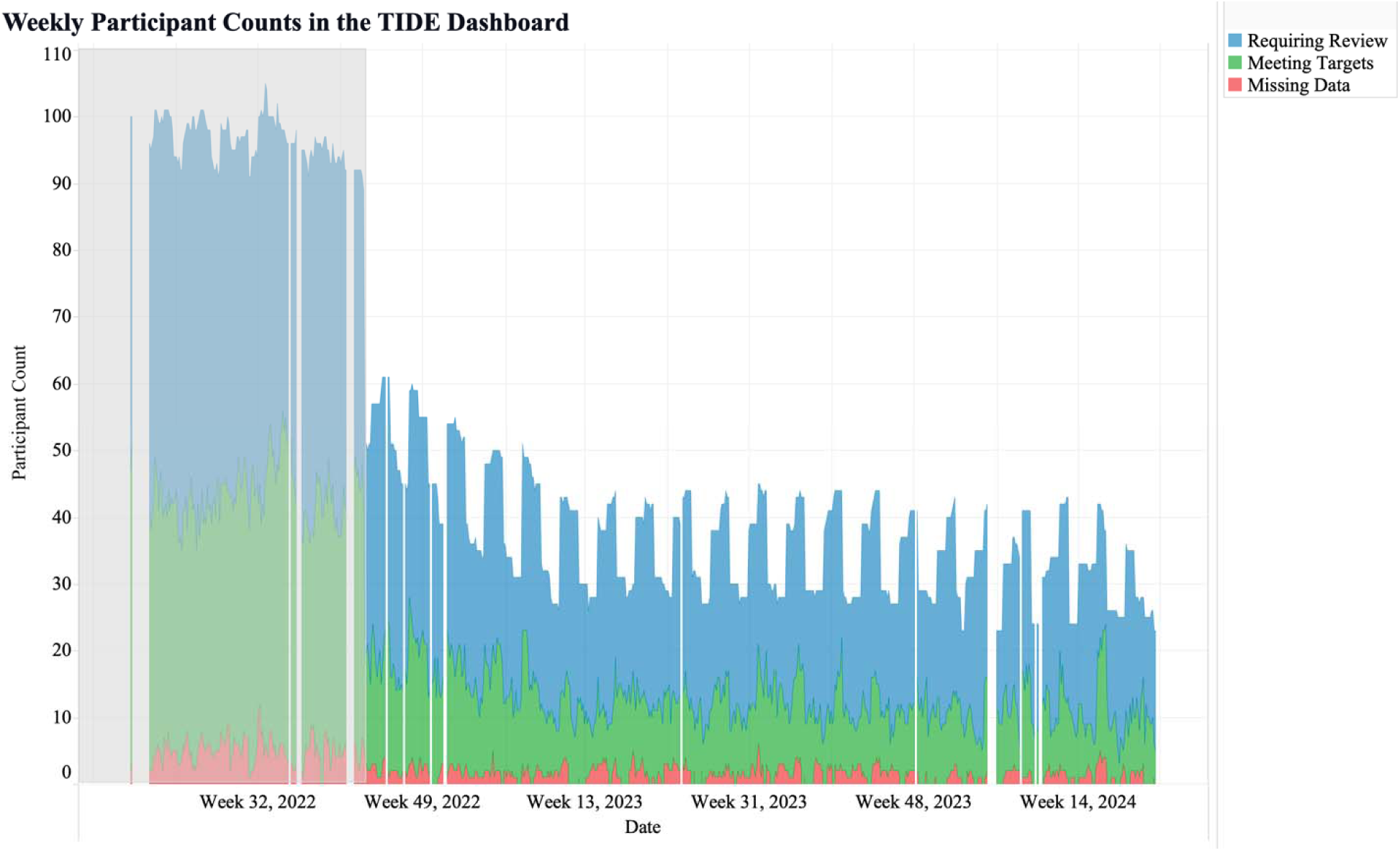
Weekly participant counts in the Timely Interventions for Diabetes Excellence (TIDE) dashboard, categorized by participants requiring review (blue), meeting targets (green), and with over 50% missing data (red). The grey area represents the duration of 4T Pilot, and the white area represents the ongoing duration of the 4T Pilot long-term follow-up program.

The second metric was the number of participants shown in TIDE categorized by each CDCES in the program. CDCES names are numbered to protect anonymity. This metric revealed significant differences in the number of participants assigned to each CDCES, as well as stability in the number of participants reviewed by each CDCES over time (Figure 2). Differences in the number of patients assigned to each CDCES can be attributed to the level of their patients’ complexity, their patients’ primary languages, and each CDCES’ time available to review participant data. This metric allowed for the monitoring of CDCES workload and identifying which CDCES team members may experience heavier participant workloads and require additional support, enabling workload balancing. For example, this metric captured the redistribution of participants from CDCES 1 to CDCES 2 in Week 12 of 2023 (Figure 2).

**Figure 2.**
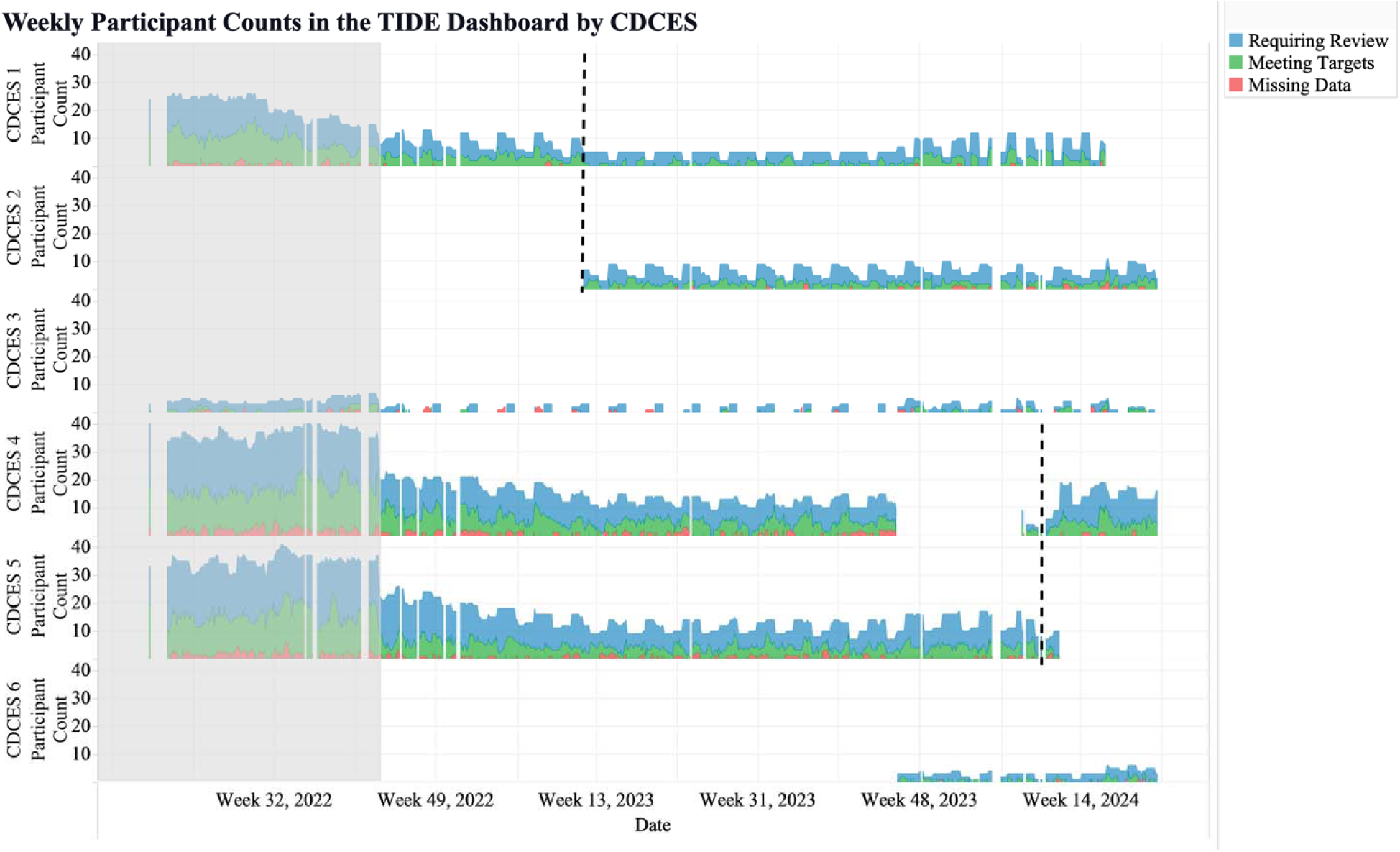
Weekly participant counts per CDCES in the TIDE dashboard, categorized by participants requiring review (blue), meeting targets (green), and with over 50% missing data (red). The dashed lines mark the transitions of participants between CDCES team members. The grey area represents the duration of 4T Pilot, and the white area represents the ongoing duration of the 4T Pilot long-term follow-up program.

Similarly, this metric showed that CDCES 5 stopped reviewing data in Week 11 of 2024, and CDCES 4 took on the review of their participants (Figure 2).

The third metric was the weekly number of participants shown in TIDE per study. This metric may help clinics with several studies view the number of participants in the RPM program for each study. Alternatively, this metric may be used for tracking the number of patients within various populations, such as the number of patients that are pediatric versus adult, within 1 year of T1D diagnosis, at potentially high risk for hypoglycemia, pregnant, non-English speaking, recently hospitalized, diagnosed with certain medical conditions or family histories, or using specific tools to manage their health. The major drop in weekly participants in 2022 corresponded to the switch to the 4T Pilot long-term follow up program, which reviewed participants monthly instead of weekly (Figure 3). The sharp decline in week 40 of 2022 occurred because all patients who had not yet reached a one-year duration in the 4T Study were moved to the long-term follow-up phase following the conclusion of 4T Pilot.

**Figure 3.**
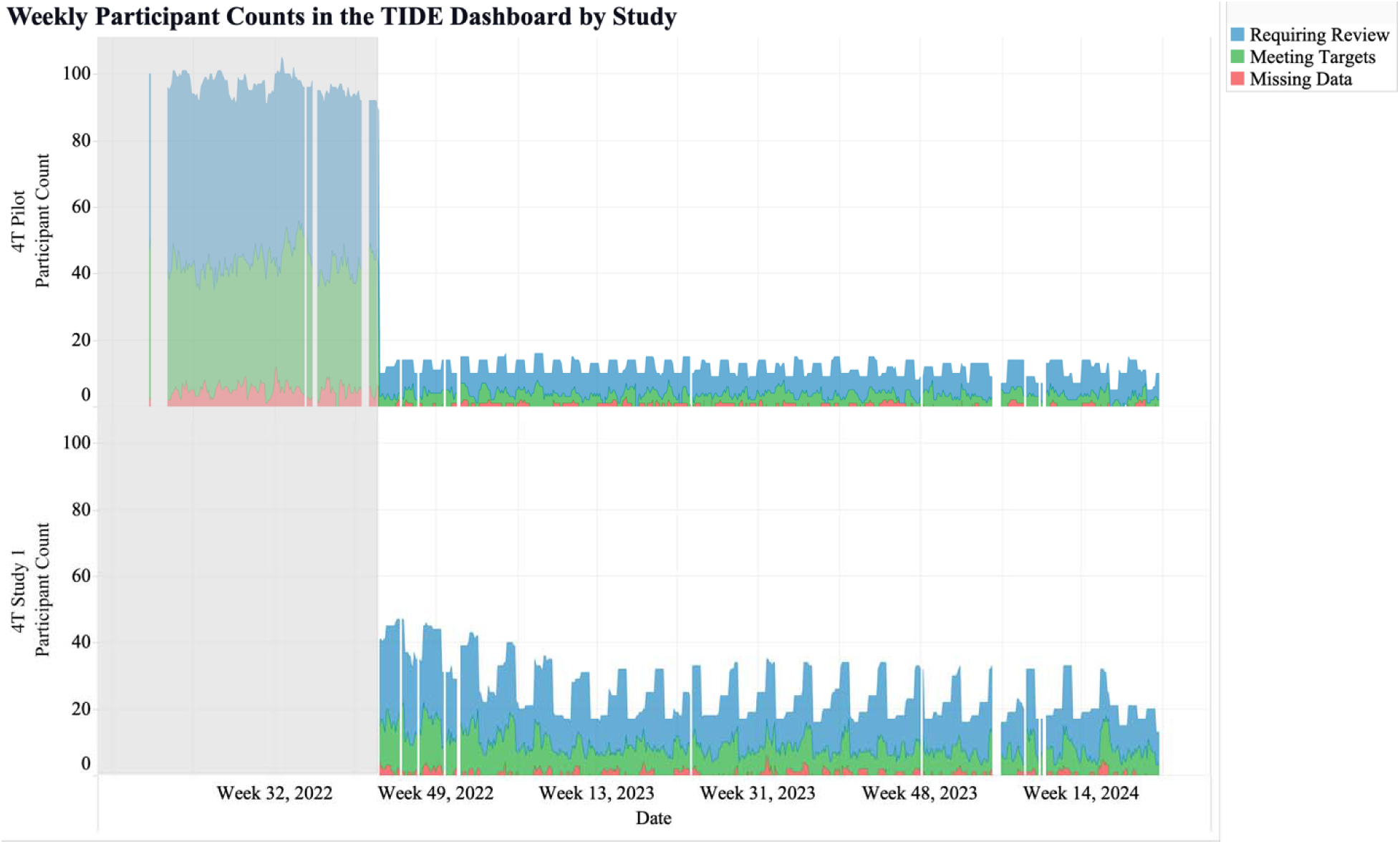
Weekly participant counts per study in the TIDE dashboard, categorized by participants requiring review (blue), meeting targets (green), and with over 50% missing data (red). The grey area represents the duration of 4T Pilot, and the white area represents the ongoing duration of the 4T Pilot long-term follow-up program.

### Metrics and Visualizations for Patient Outcomes

The fourth metric was the number of participants shown in TIDE per clinical category. This metric identified the level of variability in glucose management for each participant over time (Figure 4). This metric may help to monitor participants’ data and identify areas of improvement or concern, allowing CDCES team members to adjust care plans and interventions. For example, an increase in the number of participants missing glucose data may prompt CDCES team members to educate patients on how to improve connectivity with their CGM and how to upload glucose data.

**Figure 4.**
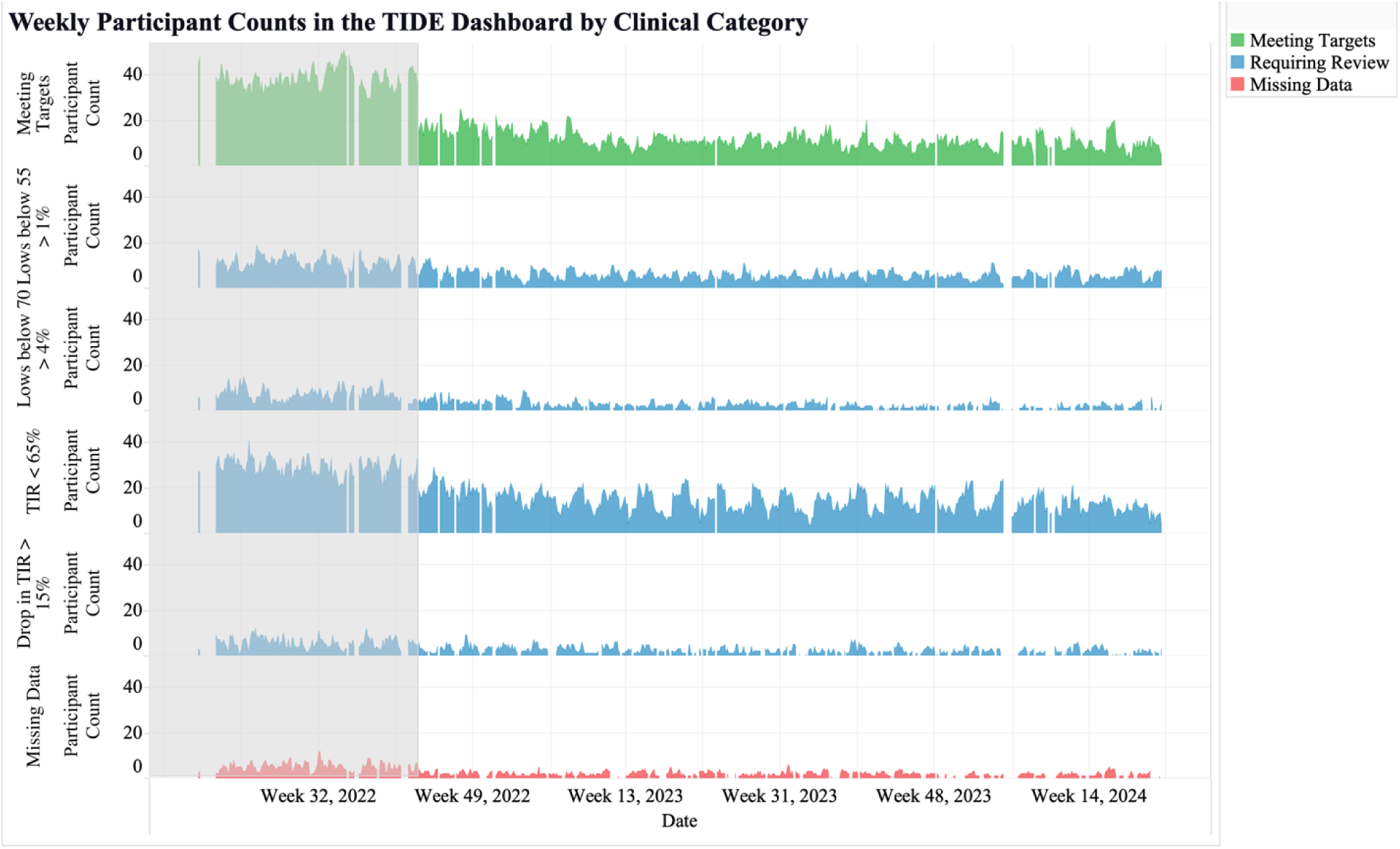
Weekly participant counts per clinical category in the TIDE dashboard, categorized by participants requiring review (blue), meeting targets (green), and with over 50% missing data (red). The grey area represents the duration of 4T Pilot, and the white area represents the ongoing duration of the 4T Pilot long-term follow-up program.

The fifth metric was the number of participants shown in TIDE per clinical category and reviewing CDCES. This metric identified the similarities and differences in glucose management for the participants reviewed by each CDCES (Figure 5). In our clinic, the metric revealed that all CDCES team members had similar proportions of participants in each clinical category (Figure 5). This metric can be used to help track how well each CDCES’ participants were meeting clinical consensus guidelines for CGM targets. The drop-off across all clinical categories before week 49 occurred because 4T Pilot ended, and not all participants chose to continue in the long-term follow-up program. Additionally, while 4T Pilot reviewed patient data weekly, the long-term follow-up program did so monthly, resulting in fewer participants appearing in the TIDE dashboard. This decline is most apparent in the green “Meeting Targets” category, as participants not meeting targets are prioritized and flagged for review over those who are meeting them.

**Figure 5.**
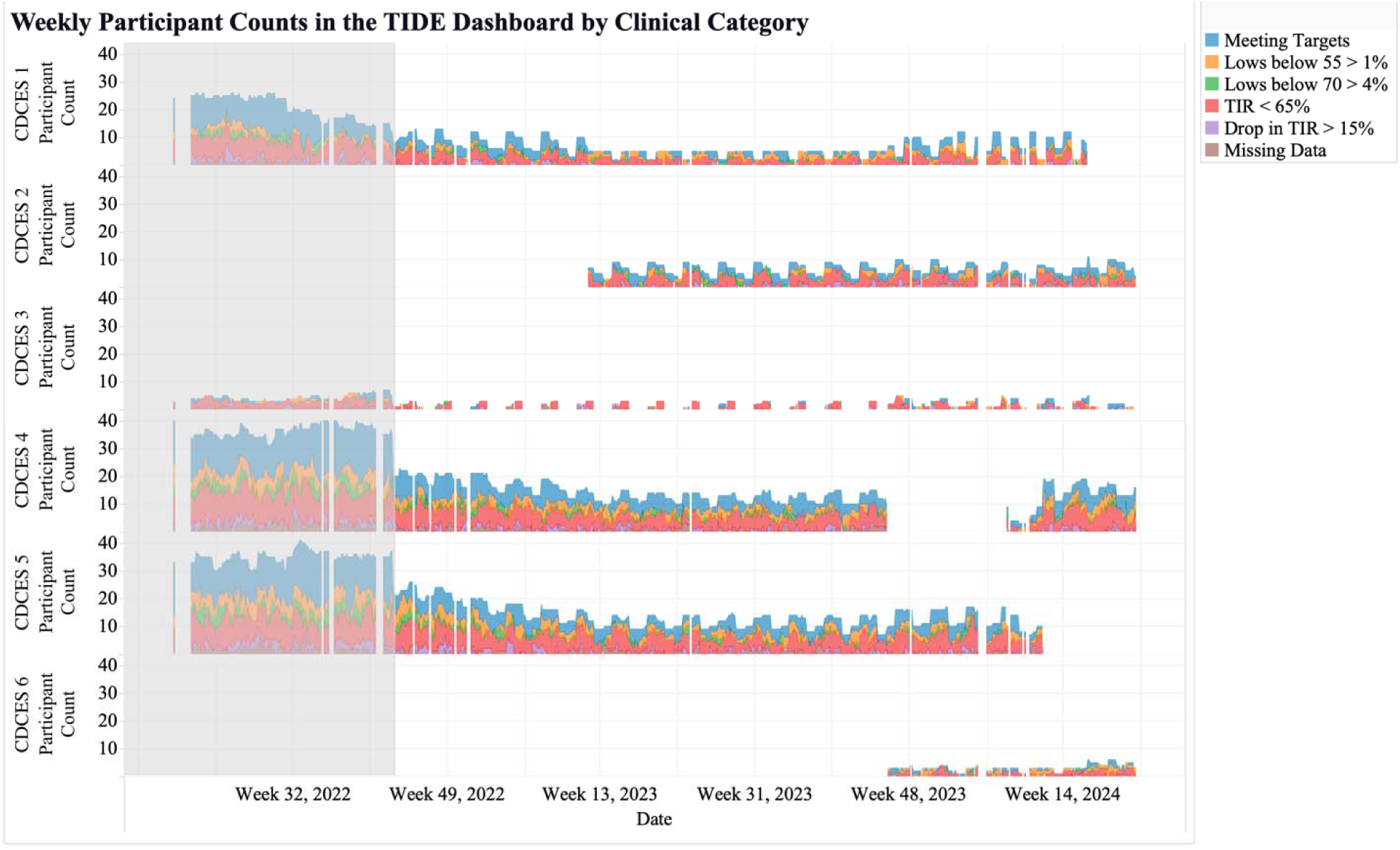
Weekly participant counts per CDCES in the TIDE dashboard, categorized by participants meeting targets (blue), spending over 1% of CGM readings below 55 mg/dL (orange), spending over 4% CGM readings below 70 mg/dL (green), spending less than 65% of CGM time in range (TIR; 70-180 mg/dL; red), experiencing over 15% drop in TIR (purple), and missing over 50% CGM data (brown). The grey area represents the duration of 4T Pilot, and the white area represents the ongoing duration of the 4T Pilot long-term follow-up program.

The sixth metric showed the weekly participant counts per study and clinical category. This metric may allow clinics to compare glucose management across studies or populations. The use of this metric allowed the 4T team to identify variation in the numbers of participants in different studies and how often the groups were reviewed by CDCES team members (Figure 6). This provided insight into how well participants in different studies met clinical consensus glycemic metrics. This metric facilitated easy comparison of health differences between groups, the difference between the outcomes of participants seen weekly versus monthly, and the clinical benefits of different study protocols.

**Figure 6.**
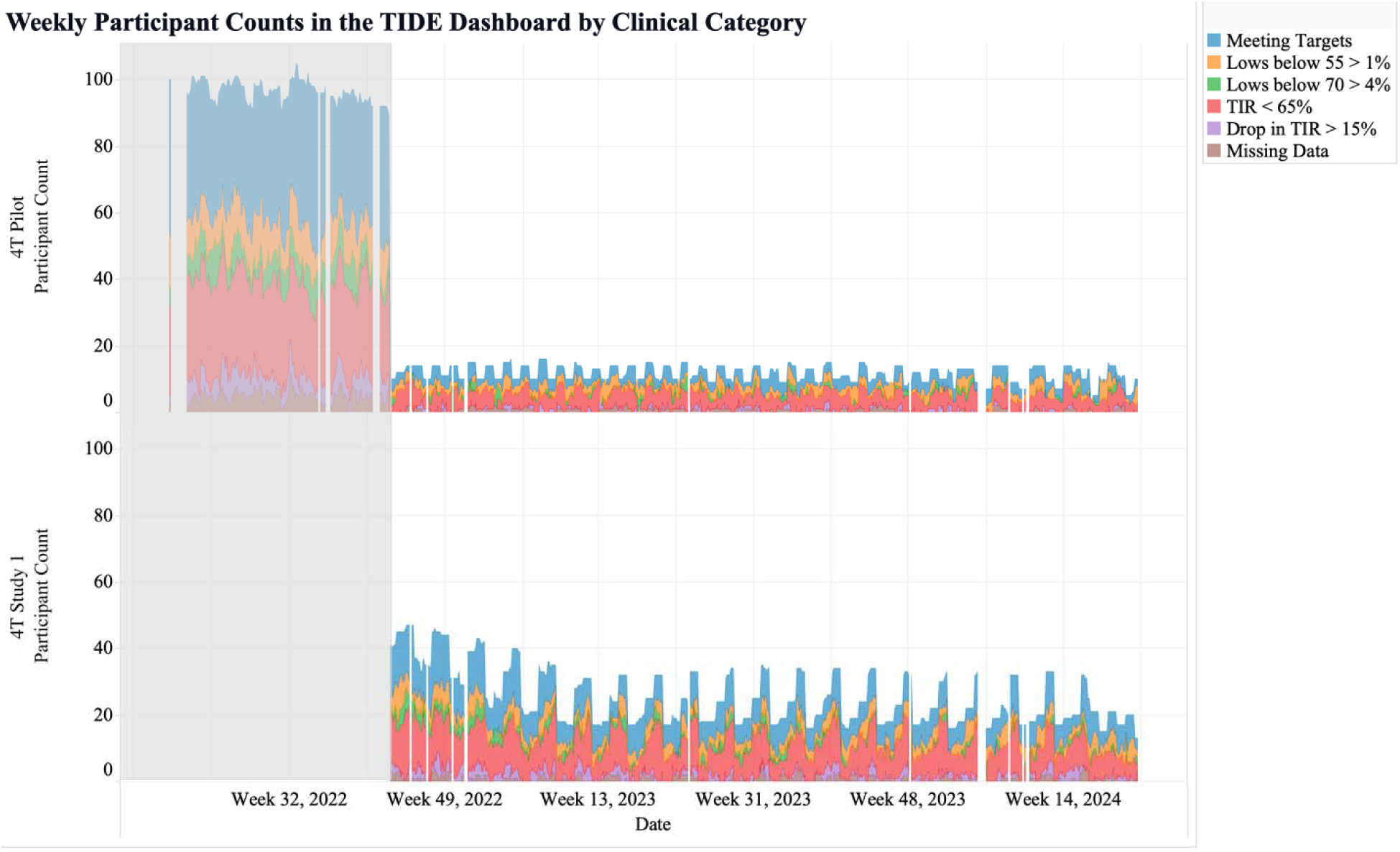
Weekly participant counts per study in the TIDE dashboard, categorized by participants meeting targets (blue), spending over 1% of CGM readings below 55 mg/dL (orange), spending over 4% CGM readings below 70 mg/dL (green), spending less than 65% of CGM time in range (TIR; 70-180 mg/dL; red), experiencing over 15% drop in TIR (purple), and missing over 50% CGM data (brown). The grey area represents the duration of 4T Pilot, and the white area represents the ongoing duration of the 4T Pilot long-term follow-up program.

### Metric and Visualization for Timeliness of Clinical Reviews

The seventh metric was originally the number of days since each participant’s most recent date of being displayed in TIDE. The patients presented were sorted from least to most recent display in TIDE and presented along with the participant’s patient ID, the CDCES that reviewed their data, and the study that participants were enrolled in. After review of this metric at a 4T leadership meeting, several alternative metrics and visualizations were proposed. The chosen visualization was a table comprising participants who have not been shown in over 20 days and ranked in the top 25% of patients with the longest time since being shown. These restrictions focused on the patients most likely to benefit from review and limit the number of patients to be reviewed by the CDCES team. In addition to showcasing the days since their last appearance in TIDE, the table included the participant’s patient ID, the CDCES responsible for them, and the study they were enrolled in. If a participant had not been flagged in the RPM program in the last 20 days, this figure allowed the CDCES team to message or schedule a visit with participants, making sure no participants were inadvertently ignored. Similarly, this metric allowed clinical research coordinators to remove certain participants from studies that they were no longer enrolled in. For example, based on this metric, research coordinators checked if the participants that have not been shown in the TIDE dashboard in over 50 days are still enrolled.

### Feedback from the Care Team

Administrative, clinical, and research feedback validated the proposed framework. Metrics were retained only if identified as helpful in understanding and visualizing clinic KPIs.

## Discussion

We developed a quantitative framework for monitoring clinical workload, patient glucose management, and timeliness of care for a whole-population T1D care model in which an algorithm analyzes CGM data to help direct care providers to patients with CGM metrics not meeting targets. Through a multi-year process of data analysis, data visualization, and feedback from the CDCES team and clinic leadership, we identified metrics that informed oversight and decision-making for the algorithm-enabled care model. This framework may be a resource for other pediatric T1D clinics seeking to optimize their RPM-based care strategies around algorithms that help direct clinician efforts.

These metrics yielded valuable insights into the RPM program and may potentially provide insights into health disparities. First, despite efforts to evenly distribute CDCES workloads, individuals meeting criteria for review were coincidentally grouped together during certain review periods, which may result in less time dedicated to individuals meeting criteria for review. This finding prompted the care team to consider redistributing subsequent enrollees. Second, the number of participants shown in TIDE per clinical category and reviewing CDCES (the fifth metric) may be used as a starting point to compare the performance of CDCES team members. Meaningful comparisons will require significant additional controls and considerations, such as patient complexity, engagement, and level of access to healthcare. Third, this metric may be used as a tool to investigate health disparities and the impact of congruence on the relationship between CDCESs and their participants. With this data, we may look into the disparity in usage of diabetes technologies, incongruence in clinician and patient perceptions of diabetes technologies, as well as how interest and feasibility of diabetes technology programs for patients with public insurance changes over time.^27^ Finally, this tool has the potential to identify positive outliers (CDCES team members whose patients achieve clinical targets) or to monitor the results of pilot interventions deployed by a subset of the CDCESs.

The ability to track clinical categories can be valuable in making decisions related to clinical efficiency and care delivery. Guided by insights from this metric, CDCES team members could stage interventions to improve glucose management or CGM wear time, compare our clinic’s TIR to other clinics, or observe how seasonal variations impact TIR.

The proposed framework is the first quantitative approach, of which we are aware, to monitor an entire RPM program’s outcomes, process metrics, and workloads. Developing metrics that drive decision making is critical, particularly in an era where algorithms can significantly influence patient care and clinical workload. In our clinic, the review of these algorithm-enabled KPIs is becoming standard practice, integral to monitoring and adjusting a care model in which algorithms reduce clinical workload.

### How to Implement KPIs and Metrics at a Clinic

The use of metrics to design KPIs can benefit large clinics or hospital systems. A large clinic or hospital might begin developing metrics by collaborating across multiple departments, potentially including internet technology specialists, data scientists, clinical research coordinators, clinicians, and quality improvement specialists. Clinics or hospitals may protect the time of or hire dedicated staff to implement the framework and help with data analysis and visualization. We recommend that the internet technology team explores several platforms to host a dashboard for visualizing their metrics. Since large clinics may face scalability or data integration issues, it could be beneficial to partner with technology vendors or research collaborators. In a collaborative effort between Stanford’s 4T team and Tidepool, a diabetes technology non-profit, Tidepool-TIDE, a new clinic-agnostic, turnkey solution available to any clinic in the United States was developed.^28^

### Limitations & Future Metrics

A limitation of the framework is the lack of data on the amount of time CDCES team members spend in the TIDE dashboard. Tableau, where TIDE is currently hosted, cannot track the amount of time that CDCES team members spend reviewing participant data. While the number of patients cared for by a CDCES is a useful proxy of workload, metrics that directly measure CDCES screen time would be ideal. Two areas for further improvement of the framework are the planned inclusion of participant reported outcomes, which are already being collected as part of the 4T program, ^11^ and metrics from physical activity trackers, such as heart rate and step count, as they become integrated into TIDE. ^13^

### Conclusion

Data from CGMs, insulin pumps, and automated insulin delivery systems is becoming increasingly important in the management of T1D. To best utilize the data, clinics increasingly turn to trusted, transparent, and user-friendly algorithms to translate patient data into insights that inform patient care. As the impact of such algorithms grows, the proposed framework may offer clinics a quantitative approach to monitor and potentially adjust the care delivery model.

## Data Availability

All data produced in the present study are available upon reasonable request to the authors.

## Acknowledgements

The authors would like to acknowledge all youth who participated in the 4T Study. We would like to thank the other members of the research team including research coordinators, clinical staff, students in the Systems Utilization Research for Stanford Medicine (SURF) group, the Quantitative Sciences Unit, the T1D Working Group in Statistics, and Informatics at Stanford Medicine Children’s Health.

## Authorship

J.K.: all (lead). J.O.F: conceptualization (equal); software (supporting). P.D: conceptualization (equal); software (supporting). D.P.Z: review and editing (supporting); methodology (supporting). D.M.M: review and editing (supporting); conceptualization (supporting); methodology (supporting). P.P: review and editing (supporting); conceptualization (supporting); methodology (supporting). R.J.: review and editing (supporting); methodology (supporting). F.K.B.: review and editing (supporting); methodology (supporting). A.A.: review and editing (supporting); methodology (supporting). D.S.: review and editing (equal); conceptualization (equal); methodology (equal).

## Funding

NIDDK - R18DK122422

Helmsley Charitable Trust (G-2002-04251-2)

ISPAD-JDRF Research Fellowship

SDRC (1P30DK 11607401)

LPCH Auxiliaries

National Science Foundation (2205084)

Stanford HAI

Stanford REDCap Platform (UL1 TR003142)

## Abbreviations

T1D: Type 1 diabetes
CGM: Continuous glucose monitor
4T: Teamwork, Targets, Technology, and Tight Control
RPM: Remote patient monitoring
TIDE: Timely Interventions for Diabetes Excellence
CDCES: Certified Diabetes Care and Education Specialist
EMR: Electronic medical record
KPI: Key performance indicator
TIR: Time in range
TBR: Time below range

