## Appendix for "A Quantitative Framework for Evaluating the Performance of Algorithm-Directed Whole-Population Remote Patient Monitoring for Type 1 Diabetes Care"

For the visualizations of the first six metrics (of seven), we placed the week of load date on the x axis and the patient count on the y axis to make a line graph. We calculated the patient count by summing up the number of unique patient ID numbers that appeared in the TIDE dashboard each week.

The first visualization (corresponding to the first metric) is composed of just the x and y axes, as well as a legend of three different patient statuses: Meeting Targets, Requiring Review, and Missing Data. The second and third visualizations feature the same legend and x and y axes, but they add another dimension. The second visualization is “stacked” with a plot for each CDCES. Each patient is assigned to a CDCES, so the patient’s ID corresponds to a CDCES name. The third visualization is also a “stacked” figure, this time with a plot for each study. Each patient ID corresponds to one study.

The fourth visualization (corresponding to the fourth metric) has the week of load date on the x axis and the patient count on the y axis with different clinical categories having their own plots, making the figure appear stacked. This figure includes a legend of patient status. The fifth visualization also has the week of load date on the x axis and the patient count on the y axis. The fifth visualization is “stacked” with a plot for each CDCES, and the legend contains each clinical category (Meeting Targets, TBR level 2 >1%, TBR level 1 > 4%, TIR < 65%, Drop in TIR > 15%, and Missing Data). The sixth visualization is the same as the fifth visualization, but each “stacked” plot now represents each study, not each CDCES.

The seventh visualization shows the number of days since a participant was shown in the RPM dashboard. As explained in the Metrics section, we display participant information only if the number of days since the participant has been shown in the TIDE dashboard exceeds 20 days and is in the highest quartile. This visualization is presented, along with additional information, in a table with four columns. The first column displays the number of days since each participant was shown in the TIDE dashboard; the second column shows the Patient ID; the third column shows the CDCES assigned to each patient; and the last column displays the study the patient enrolled in. We calculated the number of days since each participant was shown in the TIDE dashboard by using a filter to find the most recent date of each patient’s TIDE appearance. We then calculated the date difference between the date and today.
